# Efficacy of pharmacological and interventional treatment for resistant hypertension-a network meta-analysis

**DOI:** 10.1101/2023.04.21.23288951

**Authors:** Zhejia Tian, Clara Vollmer Barbosa, Hannah Lang, Johann Bauersachs, Anette Melk, Bernhard MW Schmidt

## Abstract

**Background:** Resistant hypertension is associated with a high risk of cardiovascular disease, chronic kidney disease and mortality. Yet, its management is challenging. This study aims to establish the comparative effectiveness of pharmacologic and interventional treatments by conducting a network meta-analysis.

**Methods:** MEDLINE, Cochrane Register of Controlled Trials and Web of Science Core Collection were systematically searched in March 2022. Randomized controlled trials comparing treatment options for management of resistant hypertension were included. Outcomes were blood pressure changes, measured in the office and in 24h ambulatory blood pressure measurement. We applied a frequentist random effects model to perform a network meta-analysis combining placebo medication and sham procedure as the reference comparator.

**Results:** From 4771 records, 24 studies met the inclusion criteria with 3458 included patients in total. 12 active treatment alternatives were analyzed. Among all comparators, spironolactone had the highest-ranking probability and was considered the most effective treatment to reduce office systolic blood pressure (−13.30 mmHg [−17.89; −8.72]; *P* < 0.0001) and 24h systolic blood pressure (−8.46 mmHg [−12.54; −4.38]; P < 0.0001) in patients with resistant hypertension.

**Conclusion:** Among all pharmacologic and interventional treatments, spironolactone is the most effective in reducing office and 24h systolic blood pressure in patients with resistant hypertension. More comparative trials and especially trials with long-term follow up are needed.

**Graphical Abstract:** 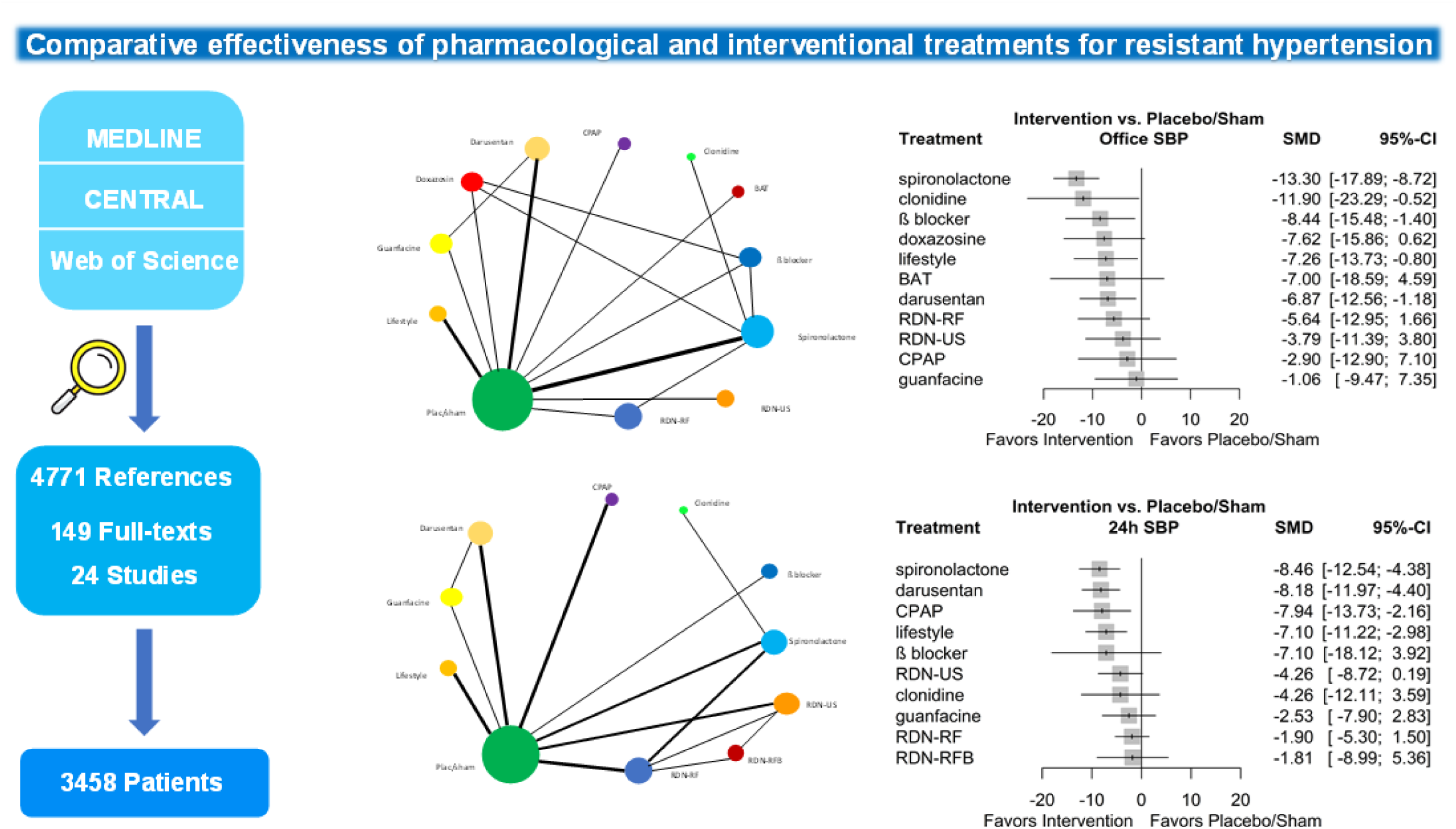

## Introduction

Hypertension remains the leading modifiable risk factor globally for cardiovascular diseases (1). There is a wealth of evidence demonstrating that lowering blood pressure (BP) reduces this risk substantially (2) (3) (4).

Resistant hypertension is defined as a blood pressure above target (>140/80 mmHg) despite the concurrent use of three different classes of antihypertensive medications at maximally tolerated doses (with one of the medications being a diuretic). For proper diagnosis, adherence to therapy should have been confirmed and pseudo-resistant hypertension and secondary causes should have been excluded (5–8). Among treated adults with hypertension, prevalent apparent treatment resistant hypertension occurs in approximately 12% to 15% of population-based reports (9) (10) (11). After applying a strict definition, the true prevalence of resistant hypertension is likely to affect <10% of treated patients (12).

Resistant hypertension accelerates hypertension-mediated organ damage, including cardiovascular and chronic kidney disease (CKD) (13) (14). Thus, there is a great demand for effective management strategies, leading to a variety of pharmacological and interventional approaches to treatment. Studies comparing different treatment options are sparse. Apart from disparities of guideline recommendations, the general approaches are to apply parallel measures, and to enhance diuretic treatment (5–8).

Network meta-analysis enables evaluation of multiple treatments simultaneously by combining direct and indirect evidence within a network of randomized controlled trials and subsequent evaluation of comparative effectiveness of different treatments (15). Hence, we accessed the comparative effectiveness, in terms of blood pressure reduction, of available pharmacologic and interventional treatments and compared it to reference treatment (placebo or sham control) in patients with resistant hypertension.

## Methods

The study was registered in the PROSPERO database (CRD42022313877). We report the study conforming to the PRISMA-NMA Extension Statement (Supplementary Table 1).

### Eligibility criteria

Trials eligible for this review were: 1) randomized controlled trials; 2) trials that enrolled patients with resistant hypertension, defined as uncontrolled hypertension despite receiving three or more antihypertensive medications, of which at least one is a diuretic; 3) trials that compared one or more treatments of interest, which defined as antihypertensive medications and interventions with approved/established efficacy to reduce blood pressure, *e.g.,* mineralocorticoid receptor antagonist (MRA), renal denervation *etc*., to each other or to placebo/sham, and 4) trials that measured changes in blood pressure as the outcomes of interest.

### Search strategy

We searched MEDLINE, Cochrane Register of Controlled Trials (CENTRAL) and Web of Science Core Collection on March 2, 2022 using following search terms: resistant hypertension AND treatment AND randomized OR randomized controlled trial without limitation of publication year.

### Study selection

Two authors independently screened the titles and abstracts for potential eligibility, and subsequently selected full-text articles. We resolved disagreements by consulting a third author. We used an online research tool (Rayyan.ai) for initial screening of titles and abstracts. For selection of full-text articles, we used a reference management software (Mendeley, Elsevier).

### Data collection and quality assessment

Two of the authors independently extracted data from included trials, with discrepancies resolved through discussion and consensus. We evaluated the quality of included trials using the Cochrane risk of bias tool (Version Aug. 2019) (16). We applied the GRADE approach to evaluate the certainty of the evidence according to Puhan et al. (17).

### Data analysis

We calculated the mean difference (MD) of BP between two treatments based on changes from baseline with standard error (SE). For outcome data only available as BP with standard deviation (SD) at baseline and endpoint we used 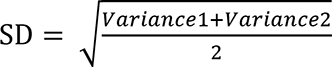 to calculate the standard deviation of change from baseline for each treatment. For outcome data presented as median with first and third quartiles we estimated the mean using the method of Luo et al. (18) and the standard deviation according to Wan et al. (19). Larger negative MD means the first treatment is more efficacious than the comparator drug.

We evaluated clinical and methodological heterogeneity as well as transitivity. Using our qualitative synthesis, we considered that participants included in our network could be randomized to any of the treatments defined in our research work (20). R version 4.2.0 was used for all calculations. First, pairwise meta-analyses for every directly compared treatment with at least 2 trials were carried out using random-effect model to evaluate the statistical heterogeneity of studies within each comparison. *metacont* function from ***meta*** package was used to calculate all pairwise treatment comparisons in multi-arm trials. A crossover trial was incorporated in our study by taking all measurements from each intervention into analysis as a parallel-trial (21). We used ***meta*** package for pairwise meta-analyses. The estimates obtained from direct comparisons are introduced as mean difference with 95% confidence interval. To detect the trials which could contribute to the statistical heterogeneity within each comparison, we further examined trial designs. Meta-regression was also applied to investigate the influence of imbalance in the covariates baseline systolic blood pressure and placebo effect.

To incorporate indirect comparisons, we then conducted random effects network meta-analyses using the ***netmeta*** package with frequentist model. For our network meta-analysis, we combined placebo and sham as the reference treatment after examining the magnitude of blood pressure change in placebo medication and sham procedure with ***meta*** package and unpaired *t* test to measure the difference. We generated the ranking probabilities of treatments according to P-scores. Heterogeneity across the network was estimated using Higgins & Thompson’s ***I^2^*** (22). We considered values above 75% as evidence of substantial heterogeneity (22). We assessed publication bias by examining funnel plots symmetry and by conducting Egger’s regression test. To further investigate inconsistency, we applied several approaches: between-designs Q statistic calculated based on a full design-by-treatment interaction random effects model (23) and τ^2^ estimated by the method of moments (24) as global approach, the net heat plot for locating inconsistency in our network and net splitting for local inconsistency. Finally, we conducted a sensitivity analysis by excluding the studies that might contribute to statistical inconsistency after checking the trial characteristics qualitatively, analyzing the heterogeneity within direct comparisons and evaluating the inconsistency throughout the network. For inconsistency and sensitivity analysis we only report results of office and 24h systolic blood pressure.

## Results

We initially identified 4771 references, 24 of which were found to be eligible for the network meta-analysis (25–48). Among them, there were two multi-arm trials and one crossover trial. The gradual selection process is outlined in Figure 1. The characteristics of included studies are summarized in Supplementary Table 2. These studies randomized a total of 3458 patients receiving 13 different treatment categories. Figure 2 illustrates the network structures.

**Figure 1.**
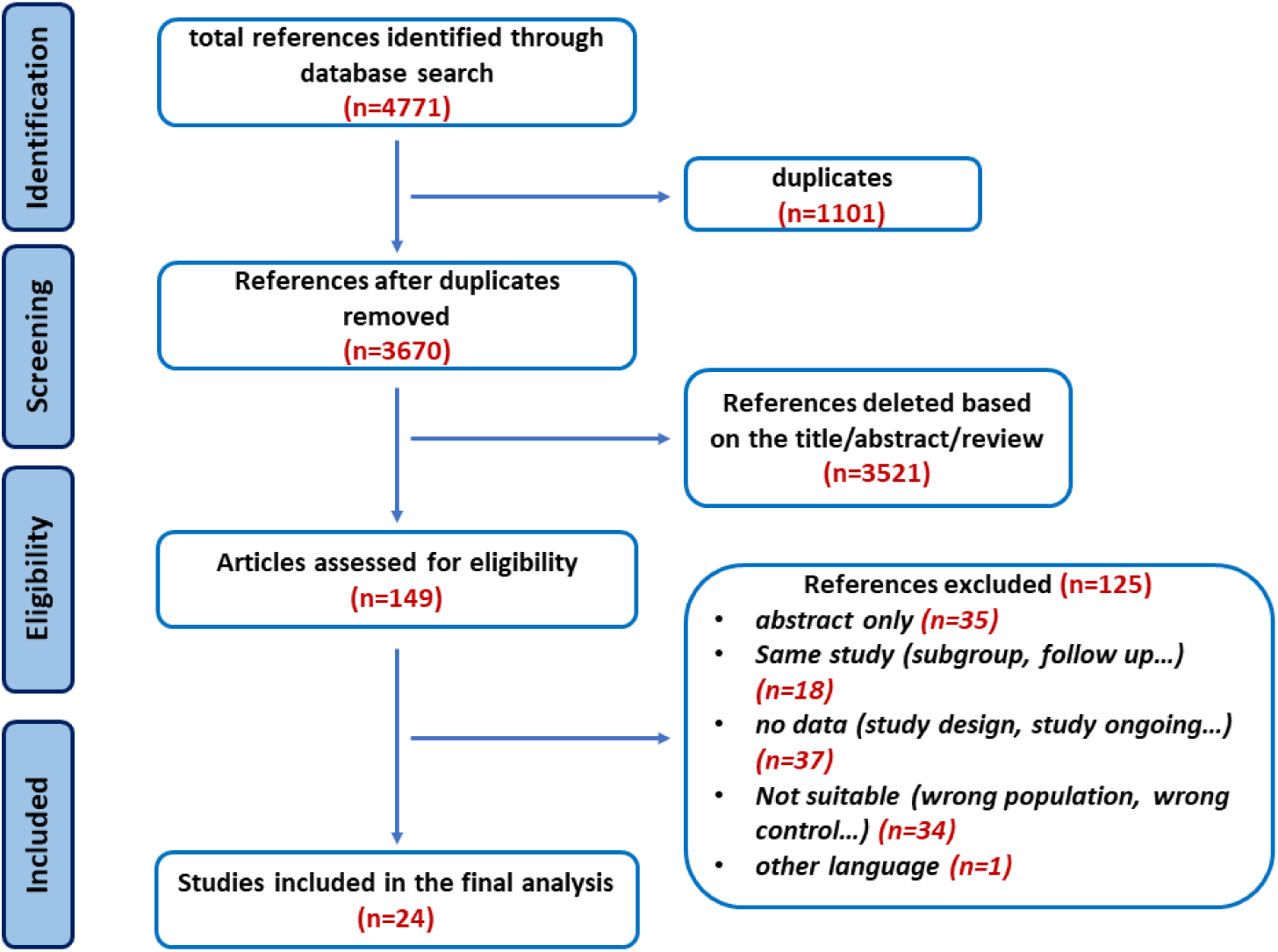
Study selection process.

**Figure 2.**
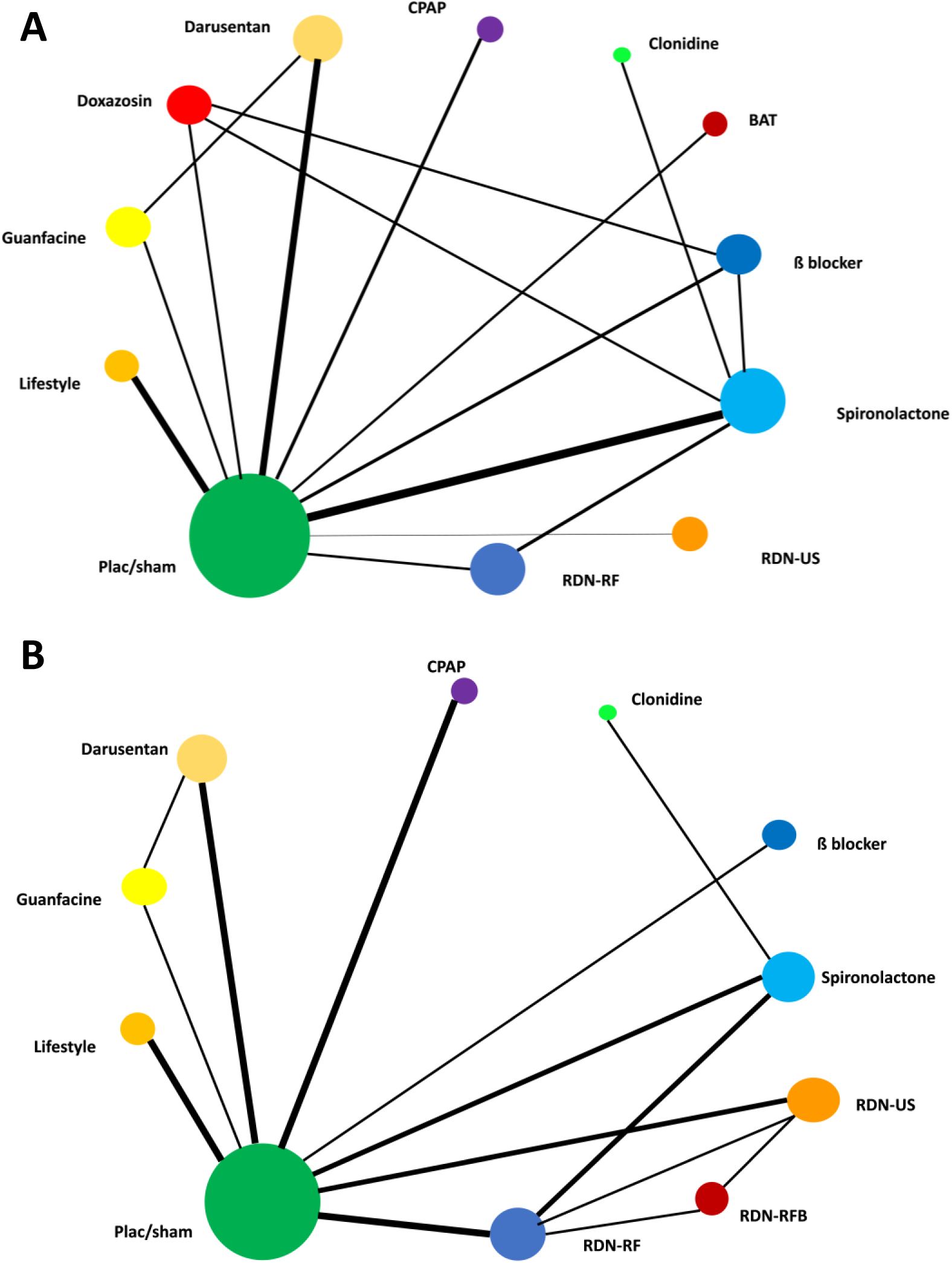
Network graph for office sBP (A) and 24h sBP (B) Each node represents one treatment. The size of the node is proportional to the number of participants randomized to that treatment. The edges represent direct comparisons. The width of the edge is proportional to the number of trials. Abbreviations: BAT baroreflex activation therapy, CPAP continuous positive airway pressure, plac/sham placebo/sham, RDN-RF standard radiofrequency-based renal denervation, RDN-RFB radiofrequency-based renal denervation with ablation distal renal arteries, RDN-US ultrasound-based renal denervation.

Supplementary Figure 1 shows the risk of bias. Most of the evidence showed moderate-to-good quality. The certainty indicators of these studies are shown in Supplementary Table 3 and 4, according to GRADE approach. The publication bias is outlined in Supplementary Figure 2 and 3

### Meta-analysis

The available direct comparisons for office sBP and 24h sBP are graphically depicted in network graphs (Figure 2). Table 1 shows the results based on direct comparisons, including the number of trials and different outcomes.

**Table 1.**
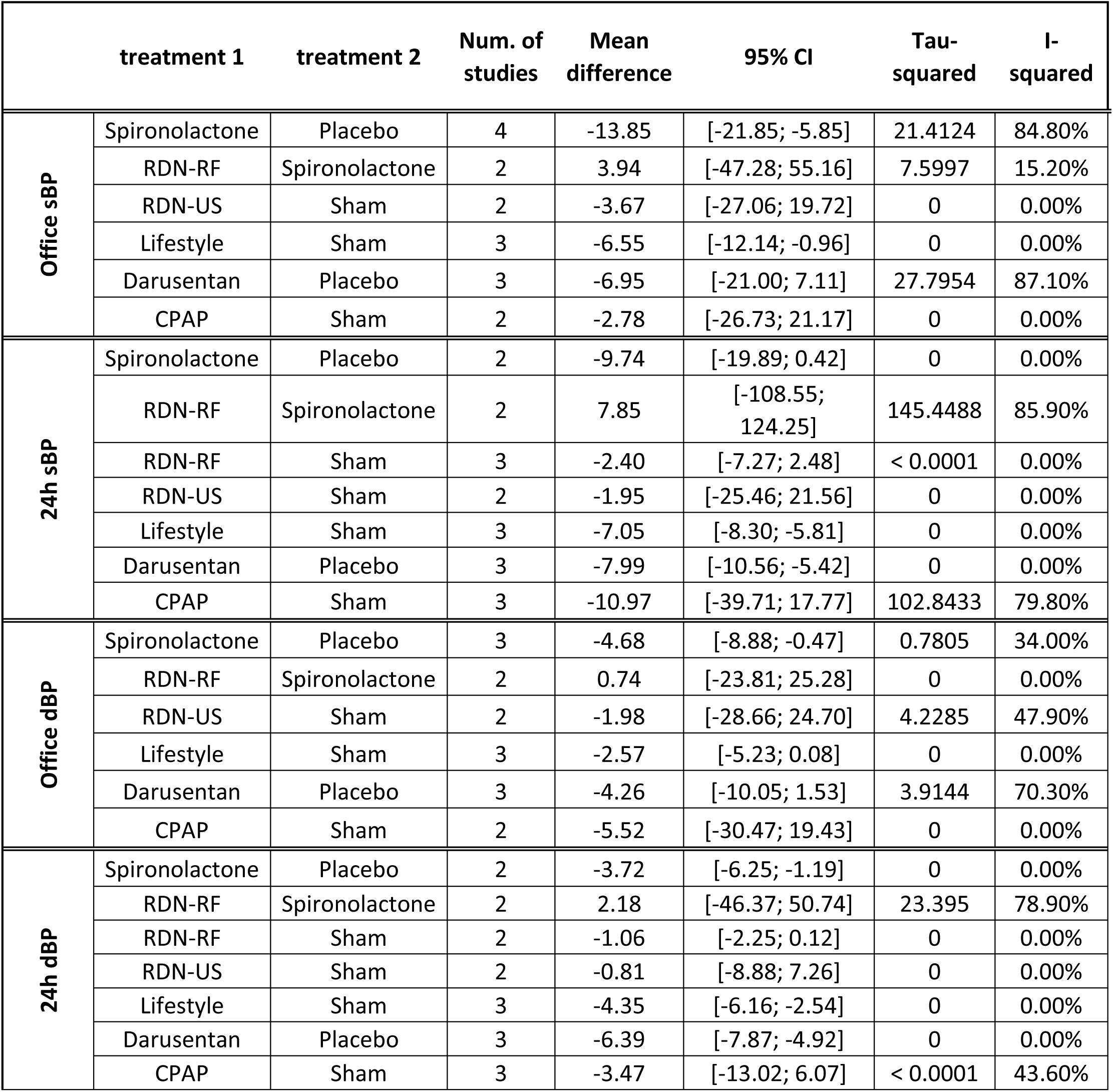
pair-wise meta-analysis based on direct comparisons. The estimates obtained from direct comparisons are presented as mean difference and 95% confidence interval. The within-comparison heterogeneity is analyzed using Higgins & Thompson’s ***I***^2^. Abbreviations: BAT baroreflex activation therapy, CPAP continuous positive airway pressure, plac/sham, placebo/sham, RDN-RF standard radiofrequency-based renal denervation, RDN-RFB radiofrequency-based renal denervation with ablation distal renal arteries, RDN-US ultrasound-based renal denervation

|  | treatment 1 | treatment 2 | Num. of studies | Mean difference | 95% CI | Tau-squared | I-squared |
| --- | --- | --- | --- | --- | --- | --- | --- |
| Office sBP | Spironolactone | Placebo | 4 | -13.85 | [-21.85; -5.85] | 21.4124 | 84.80% |
|  | RDN-RF | Spironolactone | 2 | 3.94 | [-47.28; 55.16] | 7.5997 | 15.20% |
|  | RDN-US | Sham | 2 | -3.67 | [-27.06; 19.72] | 0 | 0.00% |
|  | Lifestyle | Sham | 3 | -6.55 | [-12.14; -0.96] | 0 | 0.00% |
|  | Darusentan | Placebo | 3 | -6.95 | [-21.00; 7.11] | 27.7954 | 87.10% |
|  | CPAP | Sham | 2 | -2.78 | [-26.73; 21.17] | 0 | 0.00% |
| 24h sBP | Spironolactone | Placebo | 2 | -9.74 | [-19.89; 0.42] | 0 | 0.00% |
|  | RDN-RF | Spironolactone | 2 | 7.85 | [-108.55; 124.25] | 145.4488 | 85.90% |
|  | RDN-RF | Sham | 3 | -2.40 | [-7.27; 2.48] | < 0.0001 | 0.00% |
|  | RDN-US | Sham | 2 | -1.95 | [-25.46; 21.56] | 0 | 0.00% |
|  | Lifestyle | Sham | 3 | -7.05 | [-8.30; -5.81] | 0 | 0.00% |
|  | Darusentan | Placebo | 3 | -7.99 | [-10.56; -5.42] | 0 | 0.00% |
|  | CPAP | Sham | 3 | -10.97 | [-39.71; 17.77] | 102.8433 | 79.80% |
| Office dBP | Spironolactone | Placebo | 3 | -4.68 | [-8.88; -0.47] | 0.7805 | 34.00% |
|  | RDN-RF | Spironolactone | 2 | 0.74 | [-23.81; 25.28] | 0 | 0.00% |
|  | RDN-US | Sham | 2 | -1.98 | [-28.66; 24.70] | 4.2285 | 47.90% |
|  | Lifestyle | Sham | 3 | -2.57 | [-5.23; 0.08] | 0 | 0.00% |
|  | Darusentan | Placebo | 3 | -4.26 | [-10.05; 1.53] | 3.9144 | 70.30% |
|  | CPAP | Sham | 2 | -5.52 | [-30.47; 19.43] | 0 | 0.00% |
| 24h dBP | Spironolactone | Placebo | 2 | -3.72 | [-6.25; -1.19] | 0 | 0.00% |
|  | RDN-RF | Spironolactone | 2 | 2.18 | [-46.37; 50.74] | 23.395 | 78.90% |
|  | RDN-RF | Sham | 2 | -1.06 | [-2.25; 0.12] | 0 | 0.00% |
|  | RDN-US | Sham | 2 | -0.81 | [-8.88; 7.26] | 0 | 0.00% |
|  | Lifestyle | Sham | 3 | -4.35 | [-6.16; -2.54] | 0 | 0.00% |
|  | Darusentan | Placebo | 3 | -6.39 | [-7.87; -4.92] | 0 | 0.00% |
|  | CPAP | Sham | 3 | -3.47 | [-13.02; 6.07] | < 0.0001 | 43.60% |

### Office systolic blood pressure

Office sBP was reported in 20 trials, covering 12 treatments of interest. Spironolactone had the highest efficacy in reducing office systolic blood pressure with standardized mean difference [95% confidence interval] of −13.30 mmHg [−17.89; −8.72] (*P* < 0.0001) and had the highest probability (P-score 0.9151) of being ranked as most effective (Figure 3). Compared with placebo/sham, a significant reduction in office sBP could also be accomplished with clonidine, ß-blocker, darusentan and lifestyle management. Supplementary Figure 4 visualizes the proportion of direct and indirect evidence in our network meta-analysis. Supplementary Table 5 shows the estimates based on a frequentist network meta-analysis combining direct and indirect comparisons.

**Figure 3.**
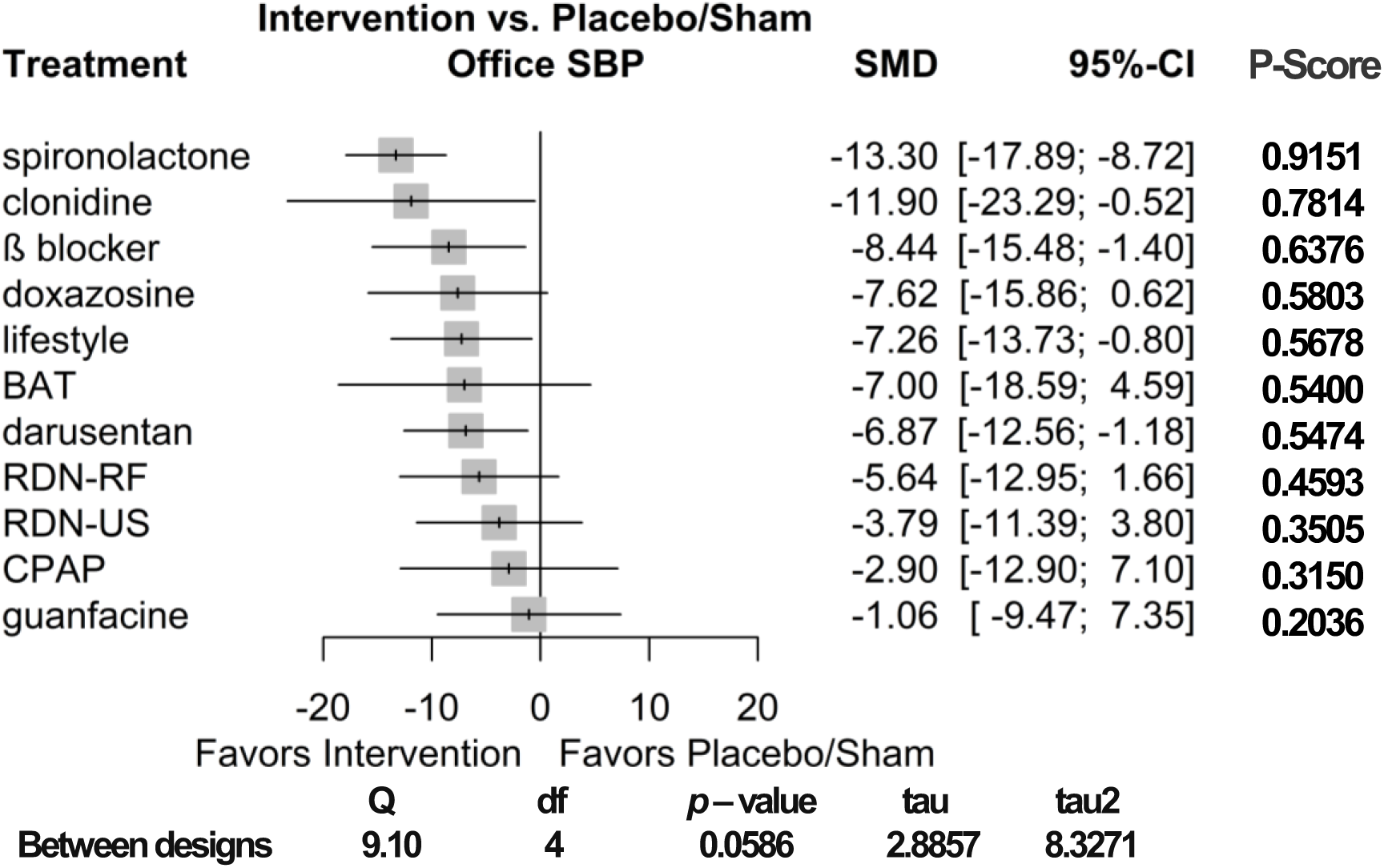
Forest plot of office sBP. Estimated effect sizes of each treatment for office sBP are presented as mean difference and 95% confidence interval. P-Score for ranking probability using frequentist model and Q statistic for inconsistency analysis are illustrated. Abbreviations: SBP systolic blood pressure, SMD standardized mean difference, BAT baroreflex activation therapy, CPAP continuous positive airway pressure, RDN-RF standard radiofrequency-based renal denervation, RDN-US ultrasound-based renal denervation.

### 24h systolic blood pressure

24h systolic blood pressure could be extracted from a total of 21 trials with 10 treatments of interest. Spironolactone was also considered the most effective treatment to reduce 24h systolic blood pressure with standardized mean difference [95% confidence interval] of −8.46 mmHg [−12.54; −4.38] (*P* < 0.0001) and the highest-ranking probability with P-score of 0.8079. Results are summarized in Figure 4, Supplementary Table 6 and Supplementary Figure 5.

**Figure 4.**
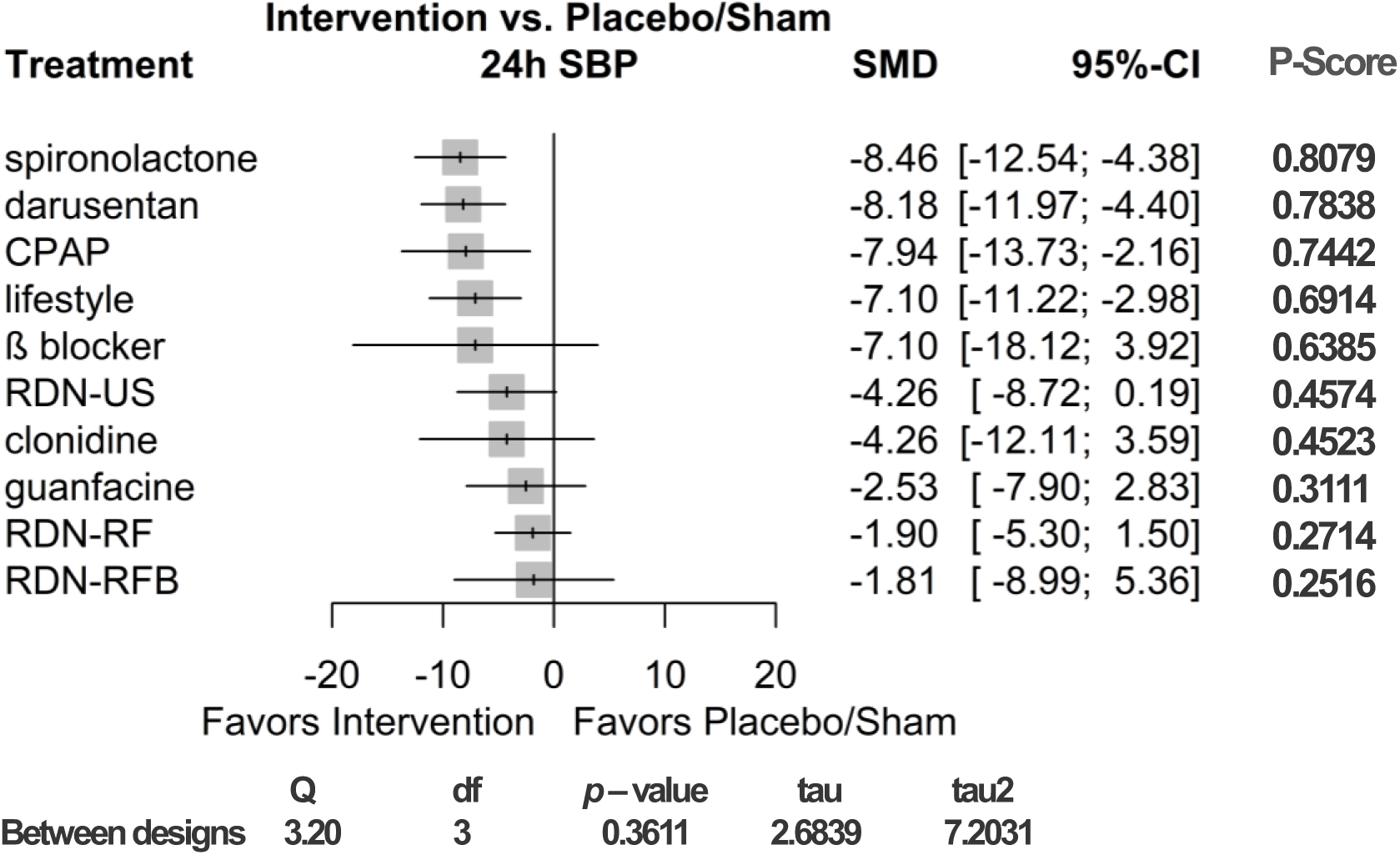
Forest plot of 24h sBP. Estimated effect sizes of each treatment for 24h sBP are presented as mean difference and 95% confidence interval. P-Score for ranking probability using frequentist model and Q statistic for inconsistency analysis are illustrated. Abbreviations: SBP systolic blood pressure, SMD standardized mean difference, CPAP continuous positive airway pressure, RDN-RF standard radiofrequency-based renal denervation, RDN-US ultrasound-based renal denervation, RDN-RFB radiofrequency-based renal denervation with ablation distal renal arteries.

### Office and 24h diastolic blood pressure

Results of office and 24h diastolic blood pressure are outlined in Figure 5. Consistent with the analyses for office sBP and 24h sBP, spironolactone lowered office dBP and 24h dBP significantly with −4.50 mmHg [−6.30; −2.70] (*P* < 0.0001) for office dBP and −3.01 mmHg [−4.92; −1.10] (*P* =0.002) for 24h dBP. According to our network meta-analysis, however, ß-blocker showed the largest effectiveness in reduction of office dBP (−5.57 mmHg [−8.01; −3.13], *P* < 0.0001), while darusentan had the highest ranking for 24h dBP (−6.50 mmHg [−8.37; −4.63], *P* < 0.0001).

**Figure 5.**
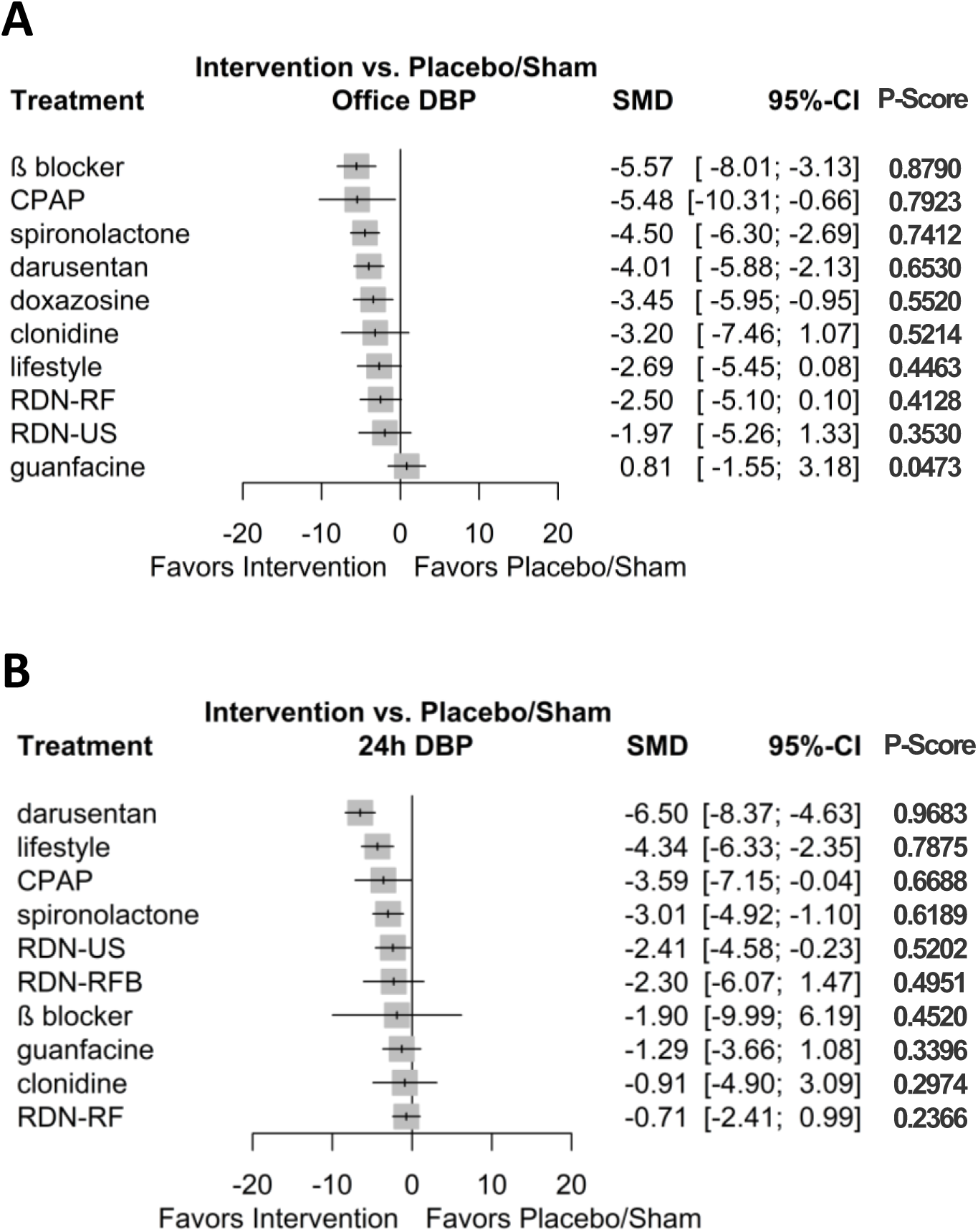
Forest plot of office dBP (A) and 24h dBP (B) Estimated effect sizes of each treatment for office dBP and 24h dBP are showed as mean difference and 95% confidence interval. Abbreviations: DBP diastolic blood pressure, SMD standardized mean difference, BAT baroreflex activation therapy, CPAP continuous positive airway pressure, RDN-RF standard radiofrequency-based renal denervation, RDN-US ultrasound-based renal denervation, RDN- RFB radiofrequency-based renal denervation with ablation distal renal arteries.

### Heterogeneity and inconsistency

Among direct comparisons, significant heterogeneity was detected in pairwise meta-analysis (Table 1). Supplementary Figure 6-12 show the heterogeneity analysis within each comparison. For accessing inconsistency in our network we implemented several methods, as mentioned above. The inconsistency analysis is outlined in Central Illustration 1 and 2 as well as in Supplementary Figure 13-16.

### Sensitivity analysis

To assess the robustness of our results, we conducted a sensitivity analysis after excluding trials that we identified as introducing statistical inconsistency into our network. The results from our sensitivity analysis, as summarized in Supplementary Figure 17 and 18, are comparable to the results of our main analysis with spironolactone as the most effective treatment, while no considerable inconsistency remained in the analysis.

### Effect of placebo/sham on blood pressure

For our network we combined placebo medication and sham procedure as the reference treatment. We explored the magnitude of blood pressure effect of placebo and sham (Supplementary Figure 19-26). We found no difference between placebo and sham effect on office sBP (t=0.7556, df=1266; P=0.45) and 24h sBP (t=0.0825, df=899; P=0.9342).

### Subgroup study

Taking the new technological development and optimization of trial designs of RDN after the SIMPLICITY HTN3-study into consideration, we performed a subgroup study of spironolactone vs. recent renal denervation studies, as illustrated in Supplementary Figure 27 and 28. Spironolactone was superior in lowering office and 24h systolic blood pressure in short-term follow up.

### Additional study

The trials on the blood pressure lowering effects of finerenone, baxdrostat and aprocitentan were published after our study selection process (49–51). We took an additional analysis to explore efficacy difference between spironolactone and the new treatments taking placebo as reference (Supplementary 29). The effect on office sBP reduction of spironolactone (−11.17 mmHg [−14.37, −7.97], P < 0.0001) and baxdrostat (−11.00 mmHg [−17.38, −4.62], P = 0.0007) was comparable, while finerenone showed less but still significant sBP reduction (−5.74 mmHg [−9.80, −1.68], P = 0.0056).

## Discussion

We conducted the first comprehensive network meta-analysis of pharmacological and interventional treatments for resistant hypertension linking the two large treatment groups by integrating placebo medication and placebo procedure into one reference comparator using trial level data. The results indicate that spironolactone is more efficacious than other treatments in reducing office and 24h systolic blood pressure with moderate-to high-quality of evidence. Although spironolactone did not show the highest probability of being ranked first for lowering diastolic blood pressure, the effect was still found to be significant. Additionally, our data reveals that clonidine, ß-blocker, lifestyle interventions and darusentan are more effective than placebo or sham in reduction of sBP.

Our findings broaden evidence-base and provide valid reference regarding the management of resistant hypertension, incorporating previous evidence from clinical trials, guidelines recommendations and several meta-analyses (5–8,52–54). If spironolactone is contraindicated or not tolerated due to e.g., high risk of hyperkalemia, alternatives such as clonidine, ß-blocker are effective alternatives. Intense multifaceted lifestyle interventions present as important principles for management of resistant hypertension. Moreover, our study suggests that in short term the pharmacologic treatments could achieve better blood pressure control compared to interventional treatments in general. Long-term effects beyond 6-12 months may be different taking into account declining compliance with medical therapy over time.

Due to the “aldosterone escape phenomenon”, a direct antagonism of aldosterone could still be needed to reach a sufficient blood pressure lowering effect, despite the reduction of aldosterone production through renin-angiotensin-aldosterone system (RAAS) Inhibition. Volume expansion and sodium retention have been well known for their contribution to resistant hypertension, which could be diminished by aldosterone-antagonism (55). According to a subsequent mechanistic analysis of PATHWAY-2, autonomous aldosterone secretion below thresholds defining primary hyperaldosteronism is prominent in patients with resistant hypertension (56).

One major safety concern in MRA therapy is hyperkalemia, especially in patients with impaired renal function and/or with concomitant RAAS blockade. A recent published retrospective cohort study indicated that initiation of an MRA for resistant hypertension substantially increased the risk of hyperkalemia (57). Especially in CKD patients, hyperkalemia may restrict the access to MRA therapy, while co-treatment with diuretics may limit hyperkalemia. Aiming to facilitate use of spironolactone among patients with resistant hypertension and advanced CKD or heart failure, randomized controlled trials with usage of potassium binder patiromer showed significantly lower serum potassium concentration in the potassium binder group (58) (59) (60). Moreover, as a well-validated CKD therapy, SGLT2 inhibitors are able to attenuate hyperkalemia, also when combined with MRA (61) (62). These findings encourage to extend the application of MRA to patients with resistant hypertension and CKD.

With the development of non-steroidal MRAs, which improve aldosterone antagonism efficacy and safety profile, we could expect a more promising pharmacologic treatment for resistant hypertension. In a post hoc analysis of finerenone in patients from FIDELITY with resistant hypertension showed a significant reduction in sBP of finerenone (49). The BLOCK-CKD trial analyzing KBP-5074 for resistant or uncontrolled hypertension among patients with advanced CKD demonstrated its effectiveness in BP reduction with a lower-than-expected risk of hyperkalemia (63). In a recently published phase 2 trial for treatment-resistant hypertension, baxdrostat, an aldosterone synthase inhibitor, reduced blood pressure substantially in a dose-related manner (50). Head-to-head trials are required for further evaluation.

Renal sympathetic denervation as a device-based approach for resistant hypertension management has been intensively studied over the past decade. On the basis of the first two SIMPLICITY HTN trials the sham-controlled SIMPLICITY HTN 3 trial failed to prove the efficacy of RDN with radiofrequency in office sBP reduction compared to sham procedure (33). As a result, recent RDN trials employed more careful control of medication use, stricter inclusion criteria and focused on new ablation technologies. The results of recent RDN trials for resistant hypertension using new technologies and with optimized trial designs were heterogeneous (31,32,35). Our subgroup study with inclusion of only recent RDN trials indicated a superiority of spironolactone over RDN. However, as demonstrated in the recently published 36-month follow-up of SIMPLICITY HTN 3 study, the BP-lowering effect of RDN seems to even increase after a longer follow-up (64), which might be valuable considering the lower compliance with pharmacological treatments in real-world settings. As no long-term studies using spironolactone are available, long-term treatment might lead to a different picture than short term studies.

Based on the encouraging results from Weber et al. and Black et al. (41,43), a phase III study of darusentan, an endothelin receptor antagonist, failed to demonstrate a significant reduction in blood pressure (42), due to an unexplained large placebo-effect. Aprocitentan, a dual endothelin receptor antagonist, which was tested in the PRECISION trial for resistant hypertension, was shown to have a significant impact on blood pressure reduction (51) and may represent a new alternative for the future.

After early-phase clinical trials, the first-generation Rheos® system was tested in the phase III Rheos Pivotal Trial, which did not meet the primary endpoint (48) (65). The efficacy of the second-generation Barostim *neo*^TM^ was so far only examined in small open-label trials (66). More data is needed to evaluate the efficacy of BAT.

Effective treatment for resistant hypertension should also involve lifestyle changes, as guidelines recommended (5,7,8). It is important to access lifestyle factors, physical as well as nutritional, that could contribute to resistant hypertension. Our network meta-analysis, in accordance to a recent meta-analysis (67), indicated that intensive lifestyle interventions are an effective option for resistant hypertension management.

Considerable inconsistency was detected in our network, which could have led to unreliable conclusion. Our sensitivity analysis confirmed our main conclusion without any inconsistent factors. In addition, renal denervation technologies were in further development and trial design was more precise after the SIMPLICITY HTN-3 trial, leaving the possibility that the earlier sham-controlled studies in our analysis underestimated the effect. Our subgroup analysis focusing on current RDN trials, though, resulted in no different conclusion with regard to the comparison between spironolactone and RDN. Moreover, as illustrated in Supplementary Figure 4-5, direct evidence obtained from included randomized controlled trials was very limited, consequently, results of our network meta-analysis should be interpreted with caution. We applied different approaches to verify the consistency assumption, which seemed to sustain in our study. At last, trials of finerenone, baxdrostat and aprocitentan were not included in our main statistical analysis, because they were published after our study selection process. Our additional analysis showed that at least baxdrostat might be as effective as spironolactone.

## Conclusions

Spironolactone is the most effective measure to reduce blood pressure in patients with resistant hypertension among all pharmacologic and interventional treatments. Clonidine and ß-blocker may also be considered as effective alternative, while intensive lifestyle interventions should be a fundamental strategy for management of resistant hypertension. With the newest data the BP-lowering efficacy of RDN is confirmed especially in long-term follow up, thus, RDN now might be considered as another treatment option. Given the obvious need for additional treatment options, the results of the ongoing development of non-steroidal MRAs, aldosterone synthase inhibitors, endothelin inhibitors and RDN therapy are eagerly awaited.

## Perspectives

Management of resistant hypertension is a multifaceted approach. Selection of adequate antihypertensive treatment for patients with resistant hypertension must consider comorbidity, risk profiles, character and preference of patients. In most circumstances, aldosterone antagonism is an important therapeutic option that might even gain more importance with the new drugs on the horizon.

Further head-to-head studies are necessary to determine whether new developing pharmacological treatments offer greater efficacy in blood pressure reduction with a better safety profile than spironolactone. Long-term follow up could provide more insights into efficacy of renal denervation versus aldosterone antagonists in reducing major cardiovascular events in the real-life setting.

## Data Availability

The datasets were derived from sources from the trial journal publications and their supplementary appendices. The data of this study will be shared on reasonable request to the corresponding author.

## Acknowledgements

No other individuals contributed to this study substantively.

## Abbreviation

sBP: systolic blood pressure
dBP: diastolic blood pressure
RAAS: renin-angiotensin-aldosterone system
sMRA: steroidal mineralocorticoid receptor antagonist
nsMRA: non-steroidal mineralocorticoid receptor antagonist
BAT: baroreflex activation therapy
RDN: renal denervation
RDN-RF: standard radiofrequency-based renal denervation
RDN-US: ultrasound-based renal denervation
RDN-RFB: radiofrequency-based renal denervation with ablation distal renal arteries

## Novelty and Relevance

The manuscript integrates all different interventions discussed for treatment of resistant hypertension in one single analysis. It shows that spironolactone is the most effective single intervention to improve blood pressure in these patients. Several other interventions are also effective and in combination might be useful for treatment of resistant hypertension. The results help physicians to find the best treatment for their patients with resistant hypertension.

